# Positive association between country-level screening rates for chlamydia and gonorrhea and self-reported diagnoses in MSM: an ecological analysis

**DOI:** 10.1101/2022.03.10.22272182

**Authors:** Vanbaelen Thibaut, Florence Eric, Kenyon Chris

## Abstract

We assessed if there was a country-level association between intensity of screening MSM for *Neisseria gonorrhoeae* and *Chlamydia trachomatis* and self-reported incidence of these infections in 2010 and 2017. Unexpectedly, we found that screening intensity was associated with a higher incidence of these infections.

**Short summary:** We found a positive association between intensity of screening for chlamydia and gonorrhoea and the self-reported incidence of these infections in MSM in European countries.

## Introduction

A number of countries in Europe and elsewhere have raised concern about increasing incidence of STIs such as *Neisseria gonorrhoeae* (NG) and *Chlamydia trachomatis* (CT)^1^. One of the strategies proposed to counter this trend has been to screen high prevalence populations such as men who have sex with men (MSM) more intensely.^1^ Even though evidence supporting such strategies is weak, numerous preexposure prophylaxis (PrEP) guidelines recommend 3-site 3-monthly screening for CT/NG in MSM taking PrEP.^2,3^ Another important limitation of this intensive screening strategy is that it has been shown to result in very high levels of macrolide and cephalosporin consumption^4^ This could aggravate the problem of antimicrobial resistance in STIs and other bacteria.^4,5^

In the absence of randomized controlled trials, other types of analyses are required to inform policies. Observational studies, for example, could assess if screening is associated with a decrease in incidence. These studies could be performed at both individual and population levels. Marcus et al. recently published such an individual level study.^3^ Using self-reported data from the European Men-Who-Have-Sex with Men Internet Survey (EMIS) they found a positive association at the individual level between screening for CT/NG and both the probability of any CT/NG diagnosis and a symptomatic CT/NG diagnosis in the previous 12 months.

In this paper we assess the population level association. Specifically, we use the same EMIS data taken from the paper by Markus et al., to assess if the incidence of CT/NG in MSM is lower in countries with more intensive screening for CT/NG ^3^.

## Methods

### Data sources

All the data used in our analysis was taken from Markus et al ^3^. This paper calculated the country-level screening and STI incidence data from 46 countries participating in the EMIS surveys of 2010 and 2017.^3^ The EMIS surveys were internet based surveys that recruited a large number of European MSM. A self-interviewing format was used to obtain information about variables such as sexual behavior, STI screening and STI diagnoses. Two waves of the study took place in 2010 and in 2017 and collected answers from more than 180,000 and 127,000 participants, respectively. The complete study methodology has been described elsewhere.^6^

#### CT/NG incidence variable

Participants were asked: “have you ever been diagnosed with gonorrhea/chlamydia or LGV?” Those who responded yes, were then asked: “when were you last diagnosed with Gonorrhea/Chlamydia or LGV?” Gonorrhea and chlamydia/LGV diagnosed in the past 24 hours, seven days, four weeks, six months and twelve months were grouped as “gonorrhea/chlamydia diagnosed in the past twelve months”. This data was used to calculate the percent of respondents per country who reported a diagnosis of CT or NG in the preceding 12 months.

#### Symptomatic CT/NG incidence variable

Participants were asked if symptoms were present at the last CT/NG test. Self-reported CT/NG diagnoses were classified as symptomatic if the diagnosis was made at the last CT/NG test and symptoms were reported at that time. The percent of respondents reporting a symptomatic episode of CT/NG per country was calculated.

#### Intensity of CT/NG screening variable

Marcus et al., constructed a new variable where being screened for CT/NG was defined as reporting a test on a urine/urethral swab or an anal swab as part of STI-testing in the previous 12 months. In addition, to distinguish screening from testing individuals with symptomatic STIs, the respondents were required to report being asymptomatic when this test was performed. The intensity of CT/NG screening per country was defined as the percent of respondents per country that reported this type of screening in the prior 12 months.

### Data analysis

Data were presented as range, median and IQR. The correlation between screening intensity and CT/NG diagnoses was tested using Spearman’s coefficient. Changes in screening intensity and STI diagnoses between the two surveys was assessed with Wilcoxon signed-rank test. The statistical analysis was performed using R version 4.0.2.

## Results

### Reported screening for CT/NG

Data was available for 40 countries. Screening intensity for CT/NG ranged from 0.4% to 22.9% (median 2.2%, IQR 1.5%-4.6%) in 2010 and from 0.8% to 36% (median 5.85%, IQR 2.55%-11.1%) in 2017. Globally, screening intensity increased between the two waves of the study (p-value = 0.001).

### Reported diagnoses of CT/NG

There were also large variations in the proportion of participants reporting a diagnosis of CT/NG in the preceding 12 months. In 2010, this proportion ranged from 1% to 10.1% (median 3.5%, IQR 2.3%-5.1%) and in 2017 from 1.1% to 16.4% (median 5%; IQR 3.3%-7.8%). This proportion also increased between the two waves of the study (p-value = 0.007).

### Reported symptomatic episodes of CT/NG

The proportion of respondents reporting a symptomatic episode of CT/NG ranged from 0% to 3.6% (median 1.25%; IQR 0.7%-1.9%) in 2010 and from 0% to 3.9% (median 1.5%; IQR 0.9%-2.23%) in 2017. This proportion increased between the two waves of the study (p-value = 0.001).

### Correlation between screening intensity and incidence of CT/NG

The intensity of screening in 2010 was positively correlated with the proportion reporting a diagnosis of CT/NG in the preceding year (rho 0.58, p-value < 0.0001). Similar results were obtained for 2017 (rho = 0.59, p-value < 0.0001; Figure 1).

**Figure 1.**
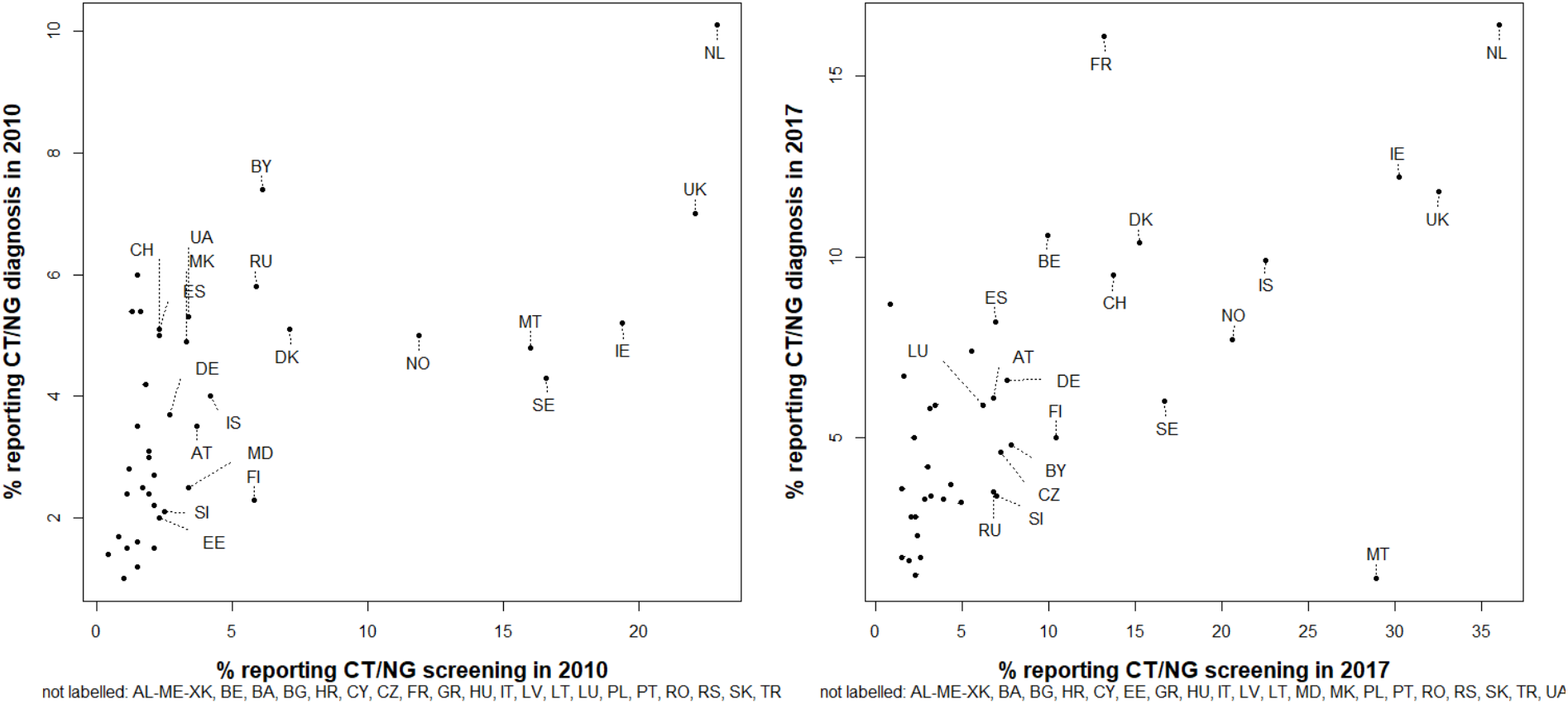
Scatter plot of country-level screening intensity for CT/NG and diagnoses of CT/NG. AL-ME-XK = Albania/Montenegro/Kosovo, AT = Austria, BY = Belarus, BE = Belgium, BA = Bosnia & Herzegovina, BG = Bulgaria, HR = Croatia, CY = Cyprus, CZ = Czech Republic, DK = Denmark, EE = Estonia, FI = Finland, FR = France, DE = Germany, GR = Greece, HU = Hungary, IS = Iceland, IE = Ireland, IT = Italy, LV = Latvia, LT = Lithuania, LU = Luxembourg, MT = Malta, MD = Moldova, NL = Netherlands, MK = North Macedonia, NO = Norway, PL = Poland, PT = Portugal, RO = Romania, RU = Russia, RS = Serbia, SK = Slovakia, SI = Slovenia, ES = Spain, SE = Sweden, CH = Switzerland, TR = Turkey, UA = Ukraine, UK = United Kingdom

### Correlation between screening intensity and symptomatic cases of CT/NG

The intensity of screening for CT/NG was positively correlated with the proportion reporting a symptomatic episode of CT/NG in the preceding year in 2010 (rho = 0.70; p-value = 0,0004) and in 2017 (rho = 0.54; p-value = 0.0003).

## Discussion

A key rationale for screening for CT/NG in MSM is to reduce the prevalence of these infections and the incidence of symptomatic infections. Our analysis did not find evidence that countries with more intensive screening had a lower incidence of CT/NG. Rather we found that screening intensity was positively associated with self-reported CT/NG diagnoses.

A likely reason for this positive association is that most CT/NG infections in MSM are asymptomatic and self-limiting.^1,7^ This means that populations that are more intensively screened are likely to have more CT/NG infections diagnosed.^7,8^ A more useful outcome measure may thus be incidence of symptomatic CT/NG. Screening intensity was, however, also not associated with a lower incidence of symptomatic CT/NG. Our study therefore does not provide support for intensive CT/NG screening in MSM.

Our study does, however, have several limitations. Firstly, there are limitations inherent to the EMIS surveys’ methodology^6,9^: (i) they are large, low-threshold online surveys, based on convenience samples, and respondents may not be representative of the entire European MSM population. (ii) they are based on self-reporting and screening/diagnoses are thus subject to recall bias, desirability bias, misinterpretation of questions and misattribution of STI symptoms/screening. (iii) They do not provide information about the type of tests used in each country, sensitivity and specificity of the tests could influence the performance of screening programs. Secondly, ecological studies are limited by the ecological inference fallacy.^10^ Thirdly we did not correct for potential confounders that could be associated with higher screening or diagnoses rates, like behavioral factors. Fourthly, PrEP was rolled out in Europe between 2010 and 2017 and this was not taken into account in our analysis.

These limitations notwithstanding, our results are commensurate with five other types of evidence.

Firstly, our results are concordant with the individual level analyses of the EMIS-2010 and 2017 surveys, which found that intensity of CT/NG screening was associated with an increased incidence of symptomatic and total CT/NG infections.^6,9,11^ Secondly, although no randomized controlled trials have been conducted in MSM, two large cluster randomized controlled trials in general populations did not find that screening for CT resulted in a decline in CT prevalence.^12,13^ Thirdly, a systematic review did not find that screening intensity in MSM was associated with a lower prevalence of CT/NG. The screening intensity evaluated in this review ranged from 3-site, 3-monthly testing to single site annual testing.^8^ Fourthly, a previous ecological study in Europe did not find that screening intensity in MSM was associated with a lower incidence of CT/NG in MSM attending sexual health clinics.^5^ Finally, modelling studies have reached various conclusions. For example, one study found that screening MSM could approximately halve the incidence of CT/NG^14^ whilst another found screening would have a minimal effect on CT/NG prevalence and result in a large increase in antimicrobial consumption.^15^

With the available evidence, we are unable to exclude the possibility that screening may be effective in MSM. Our results do however add to a growing body of evidence that suggests that screening may not be effective. A plausible reason for why screening may be infective in MSM populations with high rates of partner change is that the dense sexual networks result in a high equilibrium prevalence of CT/NG.^16^ Reinfection rates are high in this setting and even very frequent screening may have little effect on prevalence. An added concern is the arrested immunity hypothesis, which posits that early treatment of CT/NG prevents the development of an effective immune response against these pathogens which in turn results in an increased susceptibility to re-infections.^17^

The continued emergence of antimicrobial resistance in pathogens such as NG and *Mycoplasma genitalium*, particularly in core groups such as MSM^16,18^, provide additional motivation to urgently reevaluate the practice of screening MSM for CT/NG.

## Data Availability

All data produced in the present work are contained in the manuscript

https://pubmed.ncbi.nlm.nih.gov/33720969/

## Notes

**Conflicts of Interest and Source of Funding:** none declared.

### Competing Interest Statement

The authors have declared no competing interest.

### Funding Statement

This study did not receive any funding

### Author Declarations

All the data used can be obtained from the supplementary file of the following manuscript: doi:10.1371/journal.pone.0248582

